# Socio-ecological determinants of Mpox transmission risk in Uganda: mapping community-level risk and protective factors, a multi-district cross-sectional study

**DOI:** 10.64898/2026.08.12.26360257

**Authors:** Ingrid Ampaire, Richard Kabanda, Misaki Wayengera, Takudzwa Marembo, Cathy Asiimwe Tabaro, Mosoka Papa Fallah, Henry Kyobe Bosa

**Affiliations:** Ministry of Health, Kampala, Uganda; Makerere University, Kampala, Uganda; Africa Centres for Disease Control and Prevention, African Union Commission, Roosevelt Street (Old Airport), P.O. Box 3243, Addis Ababa, Ethiopia

**Keywords:** Mpox, monkeypox, socio-ecological model, risk factors, protective behaviour, Uganda, cross-sectional study, clade Ib

## Abstract

**Background:** Uganda is among the African countries most affected by the ongoing clade Ib Mpox (monkeypox) outbreak, with sustained community transmission since 2024. Effective community-level prevention depends on understanding how risk and protective factors are distributed across the social ecology. We applied a socio-ecological framework to map the determinants of Mpox transmission risk and protective practice in affected districts.

**Methods:** We conducted a multi-district, community-based cross-sectional survey of 3,960 community members aged 15–65 years across high- and low-burden districts in Uganda between February and March 2025. Participants were selected by multistage sampling and interviewed using a structured tool administered on tablets. Individual-, household-, community- and structural-level characteristics were summarised descriptively. A modified Poisson regression model with district-clustered robust standard errors was used to estimate crude and adjusted prevalence ratios (cPR, aPR) for adequate Mpox prevention and control practice; the multivariable model included 3,916 participants with complete covariate data. Analyses were performed in R version 4.5.0.

**Results:** The mean age was 32.0 years (SD 10.7); 51.9% (2,056/3,960) were female and most had secondary education (54.2%). Comorbidity burden was substantial (sexually transmitted infections 28.3%, HIV 13.9%, tuberculosis 8.2%, malnutrition 6.2%). Four in five participants (80.2%) resided in high-burden districts and 60.9% perceived themselves at risk, yet only 18.7% had comprehensive knowledge of Mpox and 53.8% reported adequate prevention practice despite near-universal message exposure (93.2%). In adjusted analysis, comprehensive knowledge (aPR 1.32, 95% CI 1.15–1.52) and message exposure (aPR 1.88, 95% CI 1.31–2.71) were by far the strongest protective determinants; several occupational, income, education and religious categories were also independently associated with practice, whereas biological comorbidities were not.

**Conclusions:** Mpox transmission risk in Uganda is concentrated in high-burden districts and overlaps with a heavy comorbidity burden. Although some social and structural factors were independently associated with protective practice, it was driven primarily by the modifiable cognitive factors of knowledge and message exposure rather than by biological characteristics. Community-level interventions that convert near-universal message reach into accurate, actionable knowledge are likely to yield the greatest protective gains.

## Introduction

Mpox (formerly monkeypox) is a viral zoonosis caused by the monkeypox virus (MPXV), an Orthopoxvirus with two genetically distinct clades. Since 2022, the disease has caused successive multi-country outbreaks, and in 2024 the emergence and rapid spread of the more virulent clade Ib sublineage from the eastern Democratic Republic of the Congo into neighbouring countries prompted the World Health Organization to declare a Public Health Emergency of International Concern and the Africa Centres for Disease Control and Prevention to declare a Public Health Emergency of Continental Security [1,2].

Uganda has become one of the most heavily affected countries in this outbreak. Following the first confirmed clade Ib cases in mid-2024, the country progressed to sustained community transmission, and by mid-2025 had reported several thousand confirmed cases and dozens of deaths, ranking among the highest case burdens on the continent [3,4]. Transmission in this setting is driven by a combination of close-contact, household and sexual exposure, occurring against a backdrop of dense urban settlement, high mobility and constrained health-system capacity [4].

Containing an outbreak of this nature requires more than biomedical countermeasures; it depends on the behaviour of communities and the social and structural conditions that shape that behaviour. The socio-ecological model offers a well-established framework for organising these influences across nested levels — individual, interpersonal, community/organisational and societal/structural — and has been widely applied to infectious-disease prevention [5,6]. Under this framework, an individual’s likelihood of acquiring or transmitting infection, and of adopting protective measures, is the product of factors operating simultaneously at each level: biological vulnerability and knowledge at the individual level; household and occupational contacts at the interpersonal level; local transmission intensity and service access at the community level; and poverty, education and normative beliefs at the structural level.

Despite this conceptual clarity, empirical mapping of how Mpox risk and protective factors are distributed across the social ecology in high-burden African settings remains limited. Most published behavioural work has focused on single domains — knowledge, attitudes or practice — without locating them within the structural and community context that determines whether protective action is feasible. Understanding this distribution is essential for designing interventions targeted to the populations and places at greatest risk.

We therefore analysed data from a large multi-district community survey conducted during Uganda’s clade Ib outbreak. Our objective was to characterise the socio-ecological determinants of Mpox transmission risk and to identify the community-level risk and protective factors associated with adequate prevention and control practice, with the aim of informing geographically and socially targeted risk-communication and community-engagement strategies.

## Methods

This study is reported in accordance with the STROBE (Strengthening the Reporting of Observational Studies in Epidemiology) guideline for cross-sectional studies; the completed checklist is provided as Supporting Information (S1 Checklist).

### Study design and setting

We conducted a descriptive, community-based cross-sectional survey in seven districts of Uganda during the clade Ib Mpox outbreak, as part of a multi-country study of the socio-ecological and behavioural drivers of Mpox [7]. Districts were purposively stratified into high-burden (Kampala, Wakiso, Mukono, Mbarara) and low-burden (Jinja, Mubende, Mityana) categories on the basis of the cumulative distribution of confirmed Mpox cases, population density, and urban mobility, allowing comparison across differing transmission intensities. Data were collected over two months, from February to March 2025.

### Participants and sampling

Eligible participants were community residents aged 15–65 years who consented to participate; adolescents aged 15–17 years were enrolled with assent in addition to guardian consent.

Participants were selected through a multistage sampling design, with sequential random selection of sub-counties, parishes, villages and households within each stratum, and one eligible respondent selected per household. A total of 3,960 participants were enrolled and contributed data to this analysis.

### Study size

The sample size was calculated using the standard formula for a single proportion in a cross-sectional survey [8]. Assuming a prevalence of comprehensive Mpox knowledge of 29% [9], 95% confidence (Z = 1.96), 90% power (Z = 1.28) and an absolute precision of 5%, the minimum requirement was 865 participants; inflation for an anticipated 10% non-response yielded 951, and a design effect reflecting Uganda’s national Mpox case burden (a multiplier of four for countries reporting 100–200 confirmed cases) gave a target of 3,804. A total of 3,960 participants were surveyed, exceeding this target.

### Variables and measurement

Data were collected by trained interviewers using a structured questionnaire administered through REDCap on tablet devices, with electronic capture into a secure database and built-in range and consistency checks. Variables were organised a priori across socio-ecological levels. Individual-level measures included age, sex, education, comprehensive knowledge of Mpox, and self-reported comorbidities (any sexually transmitted infection, HIV, tuberculosis and malnutrition). Interpersonal and household measures included marital status. Community-level measures included primary occupation and the district-level geographical risk categorisation (high-versus low-burden). Structural-level measures included daily income and religion.

Exposure to Mpox risk messaging was recorded as a binary indicator.

The primary outcome was adequate prevention and control practice, a composite binary indicator defined as endorsement of all of the following: regular hand hygiene before and after contact with objects or surfaces; willingness to consult a health professional when feeling unwell; intention to seek medical advice and self-isolate pending evaluation if Mpox were suspected; and intention to seek prompt care and testing for an affected child. Comprehensive knowledge was defined as correctly identifying three or more cardinal Mpox symptoms together with rejection of three common misconceptions.

### Bias

Several measures were taken to limit bias. Selection bias was reduced through the multistage probability sampling design within each burden stratum. Measurement bias was limited by using trained interviewers, a standardised electronic instrument with built-in validation checks, and identical procedures across districts. The potential for social-desirability and recall bias in self-reported behaviour is acknowledged and considered in the Discussion.

### Statistical analysis

Participant characteristics were summarised using frequencies and percentages; age was summarised as the mean and standard deviation (SD). Categorical characteristics were compared across burden strata using the chi-square test, and age using the Student t-test. To identify determinants of adequate prevention and control practice, we fitted a modified Poisson regression model, which yields prevalence ratios that are directly interpretable for a common binary outcome. To account for the clustering of participants within districts, standard errors were estimated using a cluster-robust (sandwich) estimator with districts as the clustering unit. Both unadjusted (crude) and adjusted prevalence ratios (cPR and aPR) with 95% confidence intervals (CI) are reported.

Multicollinearity among predictors was assessed using the variance inflation factor, and confounding and interaction were examined. The multivariable model was fitted on the 3,916 participants with complete data on all covariates (complete-case analysis); missing data on individual covariates were infrequent (fewer than 0.5% per variable). The significance threshold was set a priori at two-sided p < 0.05. Analyses were conducted in R version 4.5.0 (R Foundation for Statistical Computing, Vienna, Austria) using the package for the modified Poisson regression and cluster-robust standard errors.

### Ethical considerations

The study was conducted in accordance with the Declaration of Helsinki and Good Clinical Practice principles. Ethical approval was obtained from the Mildmay Uganda Research Ethics Committee (MUREC) (REC REF 0301-2025), and the study was registered with the Uganda National Council for Science and Technology (UNCST) (registration number HS5719ES; approved 2 April 2025). Written informed consent was obtained from all adult participants, and assent together with guardian consent was obtained for participants aged 15–17 years.

## Results

### Characteristics of study participants

A total of 3,960 participants were surveyed; the 3,916 with complete data on all model covariates were included in the multivariable analysis. The mean age was 32.0 years (SD 10.7), and slightly more than half were female (51.9%, 2,056/3,960). Most participants had attained secondary education (54.2%), while 24.0% had primary education and 14.4% had tertiary or university education. Trading or business was the most common occupation (42.1%), followed by private-sector work (24.6%), unemployment (16.1%) and farming (11.0%). The population was predominantly Christian (80.0%) and largely of low socioeconomic status, with about two-thirds earning 37,000 UGX per day or less. The comorbidity burden was considerable: 28.3% reported a sexually transmitted infection, 13.9% HIV, 8.2% tuberculosis and 6.2% malnutrition (Table 1).

**Table 1.** Socio-demographic and clinical characteristics of study participants (N = 3,960).

| Characteristic | n (%) |
| --- | --- |
| Age, years — mean (SD) | 32.0 (10.7) |
| **Sex** |  |
| Female | 2056 (51.9) |
| Male | 1903 (48.1) |
| **Marital status** |  |
| Married | 1903 (48.1) |
| Single | 1638 (41.4) |
| Separated | 244 (6.2) |
| Widow(er) | 81 (2.0) |
| Divorced | 65 (1.6) |
| Other | 29 (0.7) |
| **Education level** |  |
| No education | 248 (6.3) |
| Non-formal | 37 (0.9) |
| Primary | 951 (24.0) |
| Secondary | 2148 (54.2) |
| Tertiary / university | 571 (14.4) |
| **Occupation** |  |
| Trading / business | 1667 (42.1) |
| Private sector | 974 (24.6) |
| Unemployed | 638 (16.1) |
| Farmer | 436 (11.0) |
| Civil / public servant | 237 (6.0) |
| **Religion** |  |
| Christian | 3169 (80.0) |
| Islam | 734 (18.5) |
| Traditional worshipper | 37 (0.9) |
| Other | 16 (0.4) |
| **Daily income (UGX)** |  |
| < 3,700 | 518 (13.1) |
| 3,700 – 37,000 | 2048 (51.7) |
| 40,000 – 74,000 | 631 (15.9) |
| 77,000 – 111,000 | 127 (3.2) |
| > 111,000 | 56 (1.4) |
| No income | 571 (14.4) |
| **Comorbidities** |  |
| Any sexually transmitted infection | 1121 (28.3) |
| HIV | 552 (13.9) |
| Tuberculosis | 324 (8.2) |
| Malnutrition | 246 (6.2) |
| **District Mpox burden** |  |
| High-burden | 3175 (80.2) |
| Low-burden | 777 (19.6) |

SD, standard deviation; UGX, Ugandan shillings. Percentages are of the total sample (N = 3,960) and may not sum to 100 owing to rounding and missing values (fewer than 15 per variable).

### Distribution of Mpox risk and protective indicators across the social ecology

Mpox transmission risk was geographically concentrated: four in five participants (80.2%) resided in high-burden districts, and 60.9% perceived themselves to be personally at risk of Mpox. Protective intentions were generally high — 90.7% would seek medical advice if ill, 92.4% would seek care and testing for an affected child, and 81.6% would self-isolate when symptomatic — but consistent hand hygiene was reported by only about half (48.1% always). When combined into the composite measure, adequate prevention and control practice was present in 53.8% of participants. Notably, only 18.7% demonstrated comprehensive knowledge of Mpox despite near-universal exposure to risk messaging (93.2%) (Table 2).

**Table 2.** Distribution of Mpox risk and protective indicators across socio-ecological levels.

| Socio-ecological indicator | Category | n (%) |
| --- | --- | --- |
| **Geographical risk categorisation** | High-burden | 3175 (80.2) |
|  | Low-burden | 777 (19.6) |
| **Perceived personal risk of Mpox** | At risk | 2413 (60.9) |
|  | Not at risk | 1401 (35.4) |
| Hand hygiene before/after contact | Always | 1906 (48.1) |
|  | Often / sometimes | 1784 (45.1) |
|  | Rarely / never | 181 (4.6) |
| Would self-isolate when symptomatic | Yes | 3231 (81.6) |
| Would seek medical advice if ill | Yes | 3593 (90.7) |
| Would seek care / testing for a child | Yes | 3660 (92.4) |
| Adequate prevention & control practice | Yes | 2130 (53.8) |
| Comprehensive knowledge of Mpox | Yes | 742 (18.7) |
| Exposed to Mpox risk messaging | Yes | 3692 (93.2) |

Percentages are of the total sample ( $N = 3,960$ ); the balance of each indicator comprises the complementary category and any missing responses.

### Determinants of adequate prevention and control practice

In the adjusted model, the strongest determinants of protective practice were cognitive. Participants with comprehensive knowledge of Mpox were 32% more likely to report adequate prevention practice (aPR 1.32, 95% CI 1.15–1.52; p < 0.001), and those exposed to Mpox messaging were almost twice as likely to do so (aPR 1.88, 95% CI 1.31–2.71; p < 0.001) — by a wide margin the two largest effects in the model. Several social and structural factors were also independently associated with practice, though with smaller effects. Relative to farmers, workers in the private sector (aPR 0.76, 95% CI 0.67–0.88; p < 0.001), the unemployed (aPR 0.79, 0.69– 0.91; p < 0.001) and those in trading or business (aPR 0.86, 0.78–0.94; p = 0.001) were less likely to report adequate practice. Tertiary education (aPR 0.89, 0.80–0.98; p = 0.022), single marital status (aPR 0.92, 0.88–0.97; p = 0.003) and increasing age (aPR 0.99 per year; p = 0.037) were each associated with modestly lower practice, as were the lowest income band (aPR 0.81, 0.69–0.95; p = 0.009). Among religious groups, traditional worshippers (aPR 0.48, 0.25–0.91; p = 0.024) and adherents of other faiths (aPR 0.59, 0.49–0.71; p < 0.001) reported lower practice than Christians.

In contrast, the biological comorbidities that mark individual-level vulnerability — HIV, sexually transmitted infections, tuberculosis and malnutrition — showed no independent association with protective practice (all p > 0.05), nor did sex or district-level Mpox burden. Thus, although several structural characteristics were associated with practice, the largest and most consistent determinants by far were the modifiable cognitive factors of knowledge and message exposure (Table 3).

**Table 3.** Crude and adjusted determinants of adequate Mpox prevention and control practice (modified Poisson regression with district-clustered robust standard errors).

| Predictor | Category | cPR (95% CI) | aPR (95% CI) | p |
| --- | --- | --- | --- | --- |
| **Comprehensive knowledge** | *No (ref)* | *1.00 (ref)* | *1.00 (ref)* | — |
|  | Yes | 1.36 (1.21–1.54) | **1.32 (1.15–1.52)** | **<0.001** |
| **Mpox message exposure** | *No (ref)* | *1.00 (ref)* | *1.00 (ref)* | — |
|  | Yes | 1.97 (1.37–2.82) | **1.88 (1.31–2.71)** | **<0.001** |
| **Age (per year)** | — | 1.00 (0.99–1.00) | **0.99 (0.99–1.00)** | **0.037** |
| **Sex** | *Female (ref)* | *1.00 (ref)* | *1.00 (ref)* | — |
|  | Male | 0.98 (0.90–1.07) | 0.97 (0.88–1.06) | 0.482 |
| **Marital status** | *Married (ref)* | *1.00 (ref)* | *1.00 (ref)* | — |
|  | Single | 0.94 (0.89–0.99) | **0.92 (0.88–0.97)** | **0.003** |
|  | Separated | 0.91 (0.83–1.00) | 0.91 (0.82–1.01) | 0.071 |
|  | Divorced | 0.88 (0.69–1.12) | 0.94 (0.74–1.20) | 0.644 |
|  | Widow(er) | 0.73 (0.57–0.93) | 0.85 (0.71–1.02) | 0.077 |
|  | Other | 1.17 (0.65–2.12) | 1.18 (0.72–1.93) | 0.512 |
| **Education level** | *No education (ref)* | *1.00 (ref)* | *1.00 (ref)* | — |
|  | Non-formal | 0.81 (0.47–1.40) | 0.78 (0.42–1.45) | 0.431 |
|  | Primary | 0.97 (0.91–1.04) | 0.95 (0.87–1.04) | 0.301 |
|  | Secondary | 1.03 (0.99–1.07) | 0.95 (0.88–1.02) | 0.136 |
|  | Tertiary / university | 0.99 (0.91–1.08) | **0.89 (0.80–0.98)** | **0.022** |
| **Occupation** | *Farmer (ref)* | *1.00 (ref)* | *1.00 (ref)* | — |
|  | Trading / business | 0.88 (0.81–0.97) | **0.86 (0.78–0.94)** | **0.001** |
|  | Civil / public servant | 1.08 (0.95–1.21) | 1.00 (0.90–1.10) | 0.980 |
|  | Private sector | 0.80 (0.69–0.93) | **0.76 (0.67–0.88)** | **<0.001** |
|  | Unemployed | 0.90 (0.79–1.02) | **0.79 (0.69–0.91)** | **<0.001** |
| **Daily income (UGX)** | *No income (ref)* | *1.00 (ref)* | *1.00 (ref)* | — |
|  | < 3,700 | 0.86 (0.78–0.95) | **0.81 (0.69–0.95)** | **0.009** |
|  | 3,700 – 37,000 | 0.97 (0.91–1.03) | 0.91 (0.82–1.01) | 0.090 |
|  | 40,000 – 74,000 | 0.95 (0.85–1.06) | **0.93 (0.90–0.96)** | **<0.001** |
|  | 77,000 – 111,000 | 0.95 (0.80–1.13) | 0.99 (0.86–1.15) | 0.915 |
|  | > 111,000 | 0.93 (0.70–1.24) | 0.89 (0.61–1.29) | 0.533 |
| **Religion** | *Christian (ref)* | *1.00 (ref)* | *1.00 (ref)* | — |
|  | Islam | 0.99 (0.86–1.12) | 1.00 (0.91–1.09) | 0.939 |
|  | Traditional worshipper | 0.46 (0.25–0.83) | **0.48 (0.25–0.91)** | **0.024** |
|  | Other | 0.57 (0.46–0.71) | **0.59 (0.49–0.71)** | **<0.001** |
| **District Mpox burden** | *Low (ref)* | *1.00 (ref)* | *1.00 (ref)* | — |
|  | High | 0.95 (0.91–0.99) | 0.95 (0.90–1.01) | 0.098 |
| **HIV** | *No (ref)* | *1.00 (ref)* | *1.00 (ref)* | — |
|  | Yes | 0.99 (0.86–1.16) | 0.96 (0.88–1.06) | 0.448 |
| **Any STI** | *No (ref)* | *1.00 (ref)* | *1.00 (ref)* | — |
|  | Yes | 1.01 (0.92–1.12) | 1.02 (0.95–1.10) | 0.576 |
| **Tuberculosis** | *No (ref)* | *1.00 (ref)* | *1.00 (ref)* | — |
|  | Yes | 1.06 (0.86–1.30) | 1.06 (0.91–1.23) | 0.477 |
| **Malnutrition** | *No (ref)* | *1.00 (ref)* | *1.00 (ref)* | — |
|  | Yes | 1.04 (0.89–1.23) | 1.03 (0.89–1.20) | 0.669 |

cPR, crude (unadjusted) prevalence ratio; aPR, adjusted prevalence ratio; CI, confidence interval; ref, reference category. Estimates are from a modified Poisson regression with cluster-robust (sandwich) standard errors, districts as the clustering unit; complete-case analysis, N = 3,916. Bold indicates adjusted $p < 0.05$ . Full model output including coefficients and standard errors is provided as S1 Table.

## Discussion

In this large multi-district survey conducted during Uganda’s clade Ib Mpox outbreak, we mapped the distribution of transmission risk and protective factors across the social ecology and identified the determinants of community-level protective practice. Three findings stand out. First, transmission risk was geographically concentrated, with four in five participants living in high-burden districts and a majority perceiving themselves at risk. Second, this risk overlapped with a heavy burden of biological vulnerability — notably HIV, sexually transmitted infections and tuberculosis — clustering structural and clinical risk in the same communities. Third, and most consequential for intervention, protective practice was most strongly driven by the modifiable cognitive factors of comprehensive knowledge and message exposure, although several social and structural characteristics also showed independent, smaller associations.

The pre-eminence of knowledge and message exposure, whose effects far exceeded those of any other predictor, indicates that the social ecology shapes Mpox protection above all through the reach and quality of risk communication. Structural position was not irrelevant: relative to farmers, workers in the private sector, trade and the unemployed reported lower protective practice, as did single and older respondents, those in the lowest income band, and adherents of minority faiths. Yet these associations were modest beside the cognitive determinants, and where messaging reached people and translated into accurate knowledge, protective practice tended to follow across the social gradient. The lower practice among private-sector and other mobile occupational groups may reflect occupational mobility, reduced exposure to public-sector health messaging, or competing time pressures, and warrants targeted engagement; the lower practice among minority religious groups similarly points to a need for culturally tailored communication.

At the same time, the data reveal a critical bottleneck: near-universal message exposure (93.2%) coexisted with very low comprehensive knowledge (18.7%). Messaging is reaching communities but is not consistently being converted into the accurate, actionable understanding that drives protection. From a socio-ecological standpoint, the constraint is therefore not access to information at the structural level but the translation of that information into knowledge at the individual level. This points to a clear programmatic priority: improving the comprehensibility, cultural fit and actionability of Mpox messaging rather than simply expanding its reach.

The concentration of biological comorbidity within high-burden communities has its own implications. Although comorbidities were not associated with protective behaviour, their clustering in the populations most exposed to transmission is important for clinical preparedness, given that clade Ib is associated with more severe disease and that conditions such as HIV may worsen outcomes. Integrating Mpox prevention messaging into existing HIV, sexual-health and tuberculosis services in high-burden districts offers an efficient route to reach vulnerable groups through platforms they already use.

Taken together, these findings support a community-level prevention strategy that is geographically targeted to high-burden districts, integrated with existing chronic-disease and sexual-health services, and focused on the quality rather than merely the quantity of risk communication, while giving particular attention to mobile occupational groups and minority communities. Such an approach aligns protective effort with the socio-ecological distribution of risk that this analysis has mapped.

### Strengths and limitations

The study draws on a large, multi-district community sample spanning high- and low-burden settings, uses an explicit socio-ecological framework, and employs a modified Poisson model with district-clustered robust standard errors that appropriately accounts for the geographical structure of the data. Several limitations should be borne in mind. Because exposures and outcomes were measured concurrently in a cross-sectional design, associations cannot be interpreted causally, and the analysis characterises transmission risk and protective behaviour rather than measured transmission events. Prevention practice, comorbidities and message exposure were self-reported and may be subject to social-desirability and recall bias, which could inflate reported protective intentions. With only seven districts as clusters, district-level effects are estimated with limited precision. Despite multivariable adjustment, unmeasured factors — including specific exposure pathways and health-service access — may influence the observed associations. Finally, the findings reflect the sampled Ugandan districts during a specific outbreak phase and should be extrapolated to other settings with caution.

## Conclusions

Mpox transmission risk in Uganda is concentrated in identifiable high-burden communities and overlaps with substantial biological vulnerability. Although several social and structural characteristics were independently associated with protective practice, it was determined above all by knowledge and exposure to risk messaging rather than by biological status. The central challenge is not the reach of messaging, which is already near-universal, but its conversion into accurate, actionable knowledge. Geographically targeted, service-integrated risk communication that prioritises comprehension over coverage, with particular attention to mobile occupational groups and minority communities, is likely to deliver the greatest community-level protective benefit.

## Declarations

## Ethics approval and consent to participate

The study was approved by the Mildmay Uganda Research Ethics Committee (MUREC) (REC REF 0301-2025) and registered with the Uganda National Council for Science and Technology (UNCST) (registration number HS5719ES). All participants provided written informed consent; assent with guardian consent was obtained for participants aged 15–17 years.

## Consent for publication

Not applicable.

## Data availability

The data contain potentially identifying and sensitive participant information and cannot be made publicly available without compromising participant confidentiality. A fully de-identified minimal dataset underlying the results reported in this article is available on request, subject to ethical and governance approval, from the Mildmay Uganda Research Ethics Committee (; tel. +256 392 174236; P.O. Box 24985, Kampala, Uganda) and the corresponding author.

## Author contributions

Conceptualisation: HKB, MPF,RK; Methodology: HKB, RK, MW; Formal analysis: HKB; Investigation/data collection: IA; Data curation: AI,; Writing – original draft: IA.MT,CAT; Writing – review & editing: all authors; Supervision: HKB, MPF; Funding acquisition: MPF. All authors read and approved the final manuscript.

## Acknowledgements

The authors thank the study participants, district health teams, field interviewers and data-management staff who made this work possible, and acknowledge the contribution of local collaborators and district health offices in the participating Ugandan districts.

## Supporting information

**S1 Checklist**. STROBE checklist for cross-sectional studies.

**S1 Table**. Full regression output (crude and adjusted prevalence ratios, coefficients, standard errors, 95% CIs and p-values) for the modified Poisson model.

